# Cohort Profile: Swiss Personalized Health Network Cohort Consortium

**DOI:** 10.1101/2025.10.10.25337504

**Authors:** Murielle Bochud, Samuel El Bouzaïdi Tiali, Jan Armida, Rita Wissa, Sabine Österle, Juan Manuel Blanco, Jean Pierre Ghobril, Yves Henchoz, Valérie Pittet, Pascal Benkert, Jens Kuhle, Enrique Castelao, Martin Preisig, Carlo Chizzolini, Huldrych F. Günthard, Katharina Kusejko, Medea Imboden, Nicole Probst-Hensch, Michael Koller, Pedro Marques-Vidal, Peter Vollenweider, Menno Pruijm, Andri Rauch, Camillo Ribi, Almut Scherer, Christoph Tellenbach, Belen Ponte, Julien Vaucher, Isabel Fortier

**Affiliations:** Unisanté, University Center for Primary Care and Public Health, University of Lausanne, Switzerland; Research Institute of the McGill University Health Centre, Canada; Personalized Health Informatics Group, SIB Swiss Institute of Bioinformatics, Switzerland; Multiple Sclerosis Centre and Research Center for Clinical Neuroimmunology and Neuroscience, Departments of Biomedicine and Clinical Research, University Hospital and University of Basel, Switzerland; Department of Neurology, University Hospital and University of Basel, Switzerland; Psychiatric Epidemiology and Psychopathology Research Center, Department of Psychiatry, University Hospital and University of Lausanne, Switzerland; Clinical Immunology, Geneva University Hospital and Pathology and Immunology, School of Medicine, University of Geneva, Switzerland; Department of Infectious Diseases and Hospital Epidemiology, University Hospital Zurich, Zurich, Switzerland and Institute of Medical Virology, University of Zurich, Switzerland; Department of Epidemiology and Public Health, Swiss Tropical and Public Health Institute, Switzerland; Department of Public Health, University of Basel, Switzerland; Swiss Transplant Cohort Study, University Hospital Basel, Switzerland; Department of Medicine, Internal medicine, University Hospital and University of Lausanne, Switzerland; Department of Medicine, Service of Nephrology and Hypertension, University Hospital and university of Lausanne, Switzerland; Department of Infectious Diseases, Inselspital, University Hospital and University of Bern, Switzerland; Department of Medicine, Immunology and Allergy, University Hospital and University of Lausanne, Switzerland; Statistics Group, Swiss Clinical Quality Management Foundation, Switzerland; Nephrology and Hypertension Service, Department of Medicine, University Hospital of Geneva, Switzerland; Department of Internal Medicine and Specialties, Internal Medicine, Hospital and University of Fribourg, Switzerland

## Abstract

**Background:** Swiss cohort studies provide high-quality longitudinal data, but finding and comparing relevant studies across cohorts has historically been challenging. The Swiss Personalized Health Network Cohort Consortium (SPHN-CC) was established to address these limitations by creating the first coordinated network of Swiss cohort studies within the internationally recognized Maelstrom Research catalogue.

**Methods:** Participating cohorts were invited in 2021–2022, including longitudinal and cross-sectional studies with 1010-21 993 participants. Data collected include questionnaires, physical and cognitive assessments, administrative records, and biological samples. Variables were classified into 18 domains and 134 subdomains, and an online metadata catalogue was implemented to document study designs, explore variable content, and assess harmonization potential.

**Results:** The catalogue enables researchers to identify study-specific and harmonized variables for co-analysis. Core variables, such as age, sex/gender, anthropometrics, and medication use, are widely available, while other variables vary across cohorts. Harmonization assessments demonstrate that several key variables can be co-analyzed across multiple studies, supporting collaborative research with over 37’000 participants. A use case illustrates the potential for harmonizing and co-analyzing data across studies.

**Conclusions:** The SPHN-CC strengthens Swiss cohort research by enhancing data discoverability, supporting harmonization, and facilitating cross-cohort and international research, providing a model for more efficient use of high-value longitudinal data.

**Key features:**

- The Swiss Personalized Health Network Cohort Consortium aims to optimize the use of data and biological samples collected by publicly funded Swiss cohort studies.
- Up to now, 10 studies participated in the initiative. From 1988 to 2020 they together recruited over 50 000 participants. Recruitment remains active for six of the studies. Most cohorts are still collecting data and biological samples.
- All studies collected information from questionnaires, nine also collected biospecimens, seven performed physical measurements, two conducted cognitive assessments and two retrieved information from administrative databases at least once during the life course of the study.
- An online study and variables catalogue was developed to help researchers determine whether data collected might serve to answer the specific research questions they would like to address and, if relevant, may be harmonized and co-analyzed across studies.
- Access to the metadata catalogue is open and free.

## Why was the network set up?

Cohort studies serve as a cornerstone in clinical and public health research, providing researchers with access to longitudinal data that enables the investigation of temporal relationships between exposures and health outcomes, the progression of specific diseases, and patterns in the use of and access to healthcare services. While some cohort studies are population-based and suitable for estimating disease incidence, others are disease-specific and better suited to exploring disease trajectories, prognostic factors, and treatment outcomes. By tracking changes over time and examining relationships between health and risk factors, cohort studies offer a robust framework for uncovering insights that are difficult to attain through other study designs. Swiss cohorts, primarily publicly funded, have collected high-value data over the years. These studies track large groups of individuals over extended periods, often spanning decades, and collect health-related data across various research domains. For example, the Swiss HIV Cohort Study (SHCS) established in 1988, has yielding invaluable insights and direct benefits to the patients and general population[1,2].

Despite the important role of cohort studies, finding, accessing and understanding the data collected by these initiatives is often particularly challenging for external users. These cost- and labour-intensive studies frequently exist in isolation, with sometimes limited discoverability and substantial barriers hindering data access and/or sharing. The situation can lead to duplicated efforts and suboptimal use of the data collected. Switzerland, with its 26 cantons, has a highly decentralised health system. Each canton is largely responsible for organizing its own healthcare services, leading to considerable variation in data infrastructure, standards, and accessibility. As a result, there is no unified national health information system, and routinely collected health-related data are not easily accessible. Past efforts in establishing multi-cohort projects highlighted how administrative hurdles and time constraints can further complicate effective collaboration, emphasizing the need for dedicated resources and standardized frameworks to enhance data interoperability and research efficiency[3]. The Swiss Personalized Health Network (SPHN), launched in 2017, was established to develop nationwide infrastructure enabling the responsible use and reuse of health-related data in research[4]. As part of its efforts to assess gaps in Swiss health data interoperability, SPHN identified significant challenges related to cohort data findability, accessibility, and harmonization (i.e., processing data under a common format). This analysis highlighted the need for a structured approach to optimize cohort data utilization and foster cross-cohort research collaboration.

As part of this endeavor, SPHN supports a Cohort Consortium rallying major Swiss cohort studies. The consortium aims to enhance findability of, and accessibility to, cohorts’ data, maximize scientific value of the data collected, and leverage implementation of national and international research collaborations harmonizing and co-analysing data across studies.

As the ethical, legal and governance frameworks vary, the infrastructures differ and the data collected across studies are heterogeneous, achieving these aims is challenging. It demands the establishment of common resources optimizing the documentation and dissemination of information about study-specific characteristics and variables collected. Using the tools offered by Maelstrom Research[5], the SPHN Cohort Consortium (SPHN-CC) implemented an online metadata catalogue providing detailed information about study designs and variables collected. The catalogue allows researchers to easily grasp the data’s nature and scope without accessing the original datasets and facilitates exploring the potential to harmonize data across studies.

By systematically addressing these gaps, SPHN plays a pivotal role in strengthening Swiss cohort research, ensuring that high-value data can be more effectively leveraged for biomedical discovery and innovation and making Swiss cohorts more discoverable by the international research community.

### Who is in the network?

Participating studies were identified through calls for participation in 2021 and 2022, but membership remains open to new candidates. To be eligible, studies must be managed by a not-for-profit organisation based in Switzerland and be publicly funded. The 10 studies included to date are described in Table 1. All studies are longitudinal cohorts, except for the Swiss Health Study - Pilot Phase, which is a cross-sectional study designed to test the feasibility and acceptability of a large-scale national cohort. The number of participants recruited across studies ranges from 1010 to 21 993, with two studies including over 20 000 individuals and six still enrolling participants. Seven of the studies recruited only adults (one specifically aged 65–70 years) and three both adults and children. While five studies engaged a unique population (all participants presenting a common sampling frame and selection criteria), the others recruited different sub-populations of participants (e.g., volunteers and random sample). Finally, four studies included participants from the general population, and six included patients presenting specific diseases or conditions (e.g., rheumatic diseases, HIV, inflammatory bowel disease, multiple sclerosis, systemic lupus erythematosus). The scientific aims varied across studies with heterogeneous health outcomes of interest.

**Table 1:** Characteristics of the participating studies.

|  | Region | Participants age range | Number of sub-populations (data collection events/sub-population) | Number of participants (participants with biosamples) | Start-End year | Data sources available |
| --- | --- | --- | --- | --- | --- | --- |
| *Cardiovascular and Psychiatric arm of the Cohorte Lausannoise (CoLaus/PsyCoLaus)[18,19]* |  |  |  |  |  |  |
|  | Lausanne | 4 to 75 years | 9 (4 to 10) | 10 312 (7 769) | 2003- | Yes |
| *Lausanne cohort 65+ (Lc65+)[20,21]* |  |  |  |  |  |  |
|  | Lausanne | 65 to 70 years | 3 (6 to 16) | 4 731 (0) | 2004- | Yes |
| *Swiss Clinical Quality Management in Rheumatic Diseases (SCQM)[22]* |  |  |  |  |  |  |
|  | Whole country | Unspecified | 6 (3 to 16) | No limit | 1997- | Yes |
| *Swiss Human Immunodeficiency Virus Cohort Study (SHCS)[1]* |  |  |  |  |  |  |
|  | Whole country | 18+ years | 1 (74) | No limit | 1988- | Yes |
| *Swiss Health Study - Pilot Phase (SHeS-pp)[13]* |  |  |  |  |  |  |
|  | Whole country | 20 to 69 years | 3 (1) | 1 349 (780) | 2020-2021 | Yes |
| *Swiss Inflammatory Bowel Disease Cohort Study (SIBDCS)[23]* |  |  |  |  |  |  |
|  | Whole country | 7 to 89 years | 1 (16) | No limit | 2006- | Yes |
| *Swiss Kidney Project on Genes in Hypertension (SKIPOGH)[24]* |  |  |  |  |  |  |
|  | Lausanne, Geneva, Bern | 18+ years | 1 (2) | 1 128 (1 128) | 2009-2016 | Yes |
| *The Swiss Multiple Sclerosis Cohort-Study (SMSC)[25]* |  |  |  |  |  |  |
|  | Aarau, Basel, Berne, Geneva, Lausanne, Lugano, St. Gallen, Zürich | 18+ years | 1 (21) | No limit | 2010- | Yes |
| *Swiss Systemic Lupus Erythematosus Cohort Study (SSCS)[26]* |  |  |  |  |  |  |
|  | Basel, Bern, Geneva, Vaud, Schaffhausen, St. Gallen, Valais, Zürich and private practices | 18+ years | 4 (14 to 24) | No limit | 2007- | Yes |
| *Swiss Transplant Cohort Study (STCS)[27,28]* |  |  |  |  |  |  |
|  | Whole country | 18+ years | 1 (17) | No limit | 2008- | Yes |

## How often have participants been followed up?

Participant enrolment occurred between 1988 and 2020. Two studies launched recruitment before 2000, six between 2000 and 2009, and two after 2010. The frequency and profiles of the data collection events (DCEs) vary extensively across studies. Some followed predefined patterns of follow-up (e.g., every six months), whereas in others, the DCEs varied over time. As of today, eight of the 10 studies are still collecting data (see Table 1).

## What has been measured?

All participating studies collected information from questionnaires (10 studies), seven performed physical measurements, two conducted cognitive assessments, and two retrieved information from administrative databases at least once during the life course of the study. Nine studies also collected biological samples (nine blood, four urines, two tissues or viable cells, and one cerebrospinal fluid).

Based on the data dictionaries provided by the studies, for each DCE and subpopulation of participants, the variables collected were classified into 18 domains (e.g., sociodemographic characteristics or lifestyle and behaviours) and 134 subdomains of information (e.g., participant age or tobacco use). As a result, it is possible to explore the information content easily. All participating studies collected information about age, sex/gender, anthropometric measures, and medication intake. Nine studies collected information about education, tobacco consumption, pregnancy and birth, surgical interventions, and biochemistry measurements. Some information, on the other hand, such as life course development, misbehaviour and criminality, and transportation, is collected by only one study. For example, Figure 1 provides an overview of the subdomains of information collected by studies for three domains: socio-demographic and economic characteristics, lifestyle and behaviours, and diseases (ICD codes). Figure 2 outlines the specific information collected over time by the Lausanne cohort 65+ for the same three domains. More comprehensive information is provided in the online catalogue (https://www.maelstrom-research.org/network/sphn-cc), including a description of the specific variables collected (variable name, label, and units or specific categories).

**Figure 1.**
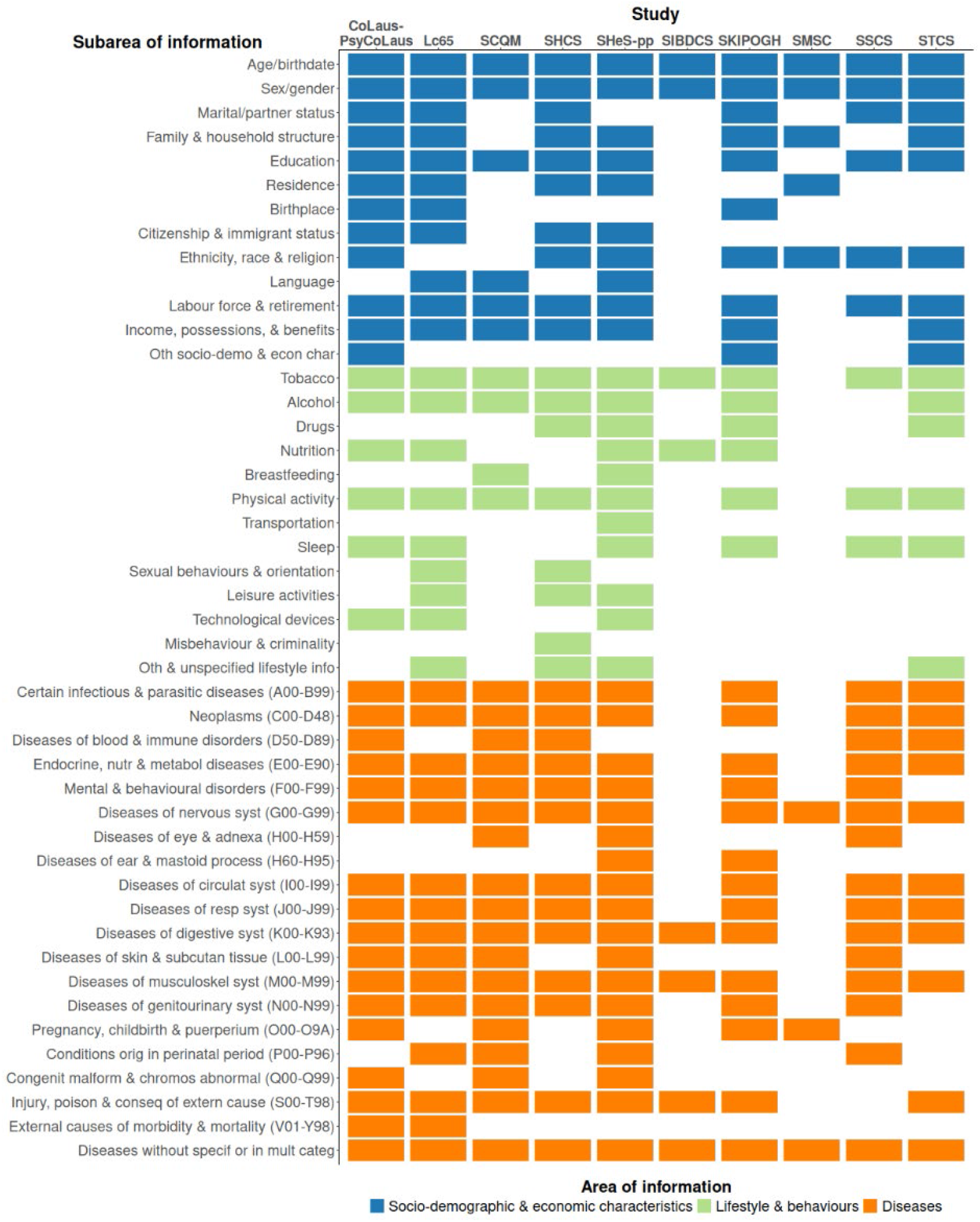
Subdomains of information collected by participating studies for the domains: socio-demographic and economic characteristics, lifestyle and behaviours and diseases (ICD10 codes).

**Figure 2.**
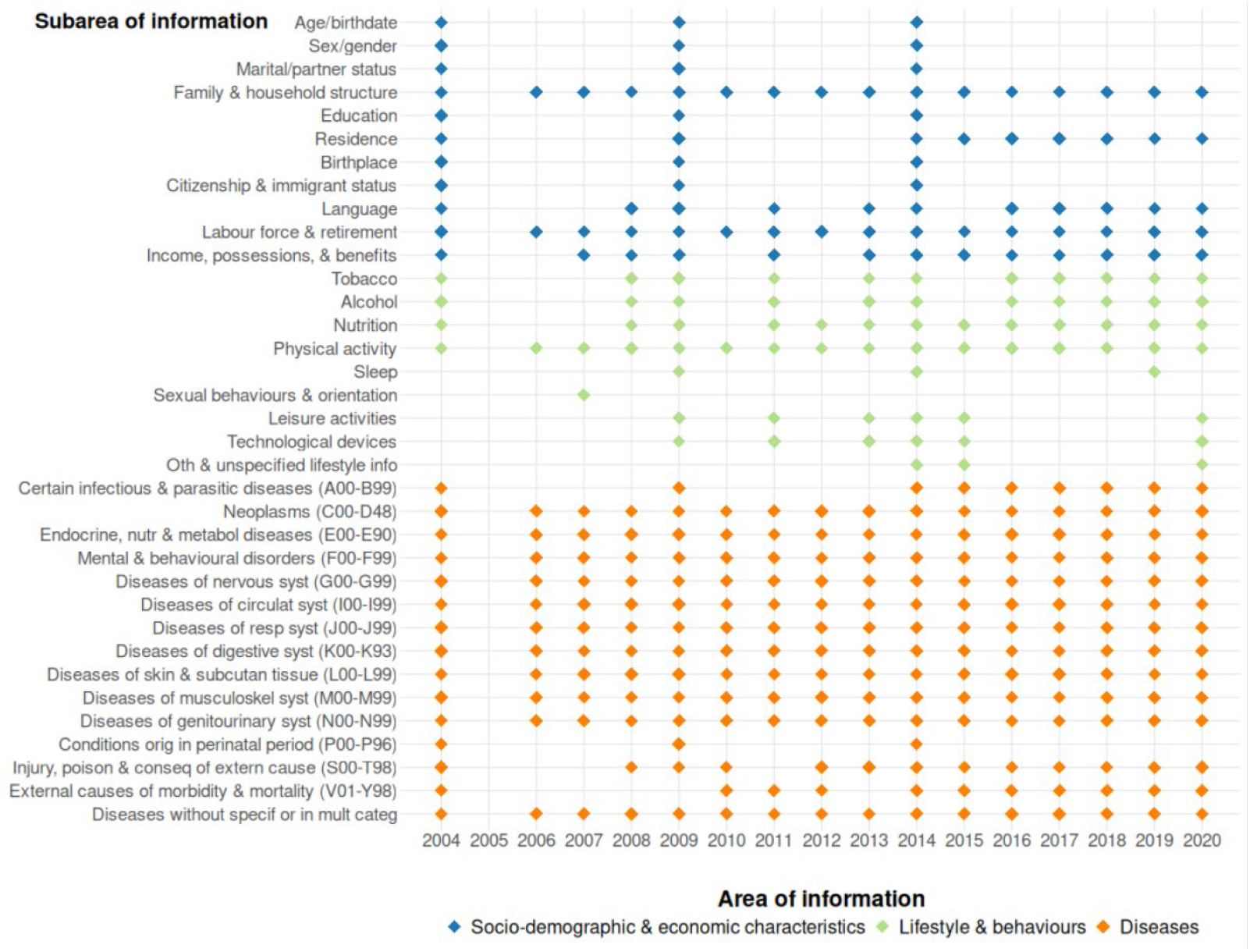
Subdomains of information collected by the Lausanne cohort 65+ through time for the domains: socio-demographic and economic characteristics, lifestyle and behaviours and diseases (ICD10 codes).

## What the metadata cataloguing and harmonization process found?

Studies participating in the SPHN-CC assembled an extensive collection of data and samples that could be instrumental in supporting a broad range of research initiatives. The study and variables catalogue implemented by the consortium allows leveraging the use of these scientific resources. The SPHN-CC catalogue helps investigators determine whether specific data of interest are available to them. It also helps identify data that can be harmonized and co-analyzed across the network studies or other studies included in the Maelstrom catalogue to answer specific research questions.

The catalogue contains comprehensive information about individual studies (e.g., study characteristics, profiles of subpopulations of participants, timing of data collection events, etc.) and content of the variables collected (over 134 000 variables). A user-friendly search engine facilitates data discovery. For example, the catalogue can help identify the study that collected information about fatigue, specifically using the Chalder Fatigue Scale[6] (CoLaus|PsyCoLaus) or those collecting anthropometric measures and information about nutrition and physical activity concurrently (CoLaus|PsyCoLaus, Lc65+, SHeS-pp and SKIPOGH).

Investigators may also be interested in asking for access, harmonizing and co-analyzing data across studies. The catalogue allows exploring the potential to use collected data to generate the harmonized variables required to support co-analysis. As the collected variables are documented and easily searchable, it is possible to evaluate if a study can (or cannot) generate a defined variable. For example, if we define weight as measured or self-reported (units= kilograms), nine of the studies can be co-analyzed, however, if we define weight as measured (units = kilograms), i.e. excluding self-reported weight, then three studies can be co-analyzed. Supplementary material 1 (Harmonization potential across partner studies for a selected ensemble of defined harmonized variables at baseline) provides a use case exploring the potential to harmonize specific data items across studies (details are also available on Maelstrom Research’s website in the SPHN Harmonization Initiative: https://www.maelstrom-research.org/study/sphn-hi). A list of 26 commonly used variables was defined, and the potential to generate each of these variables, as defined, is evaluated based on the study-specific variables collected at baseline (26 variables across 10 studies, leading to 260 harmonization estimated statuses). The harmonization status is considered ‘‘complete’’ when study-specific variables can be used to generate the defined core variable, ‘‘partial’’ when it is possible but involves a loss of information, and ‘‘impossible’’ when the core variable cannot be generated either because the study-specific variable’s definition is incompatible with the defined variable for harmonization, or because the information is simply not collected for this study at baseline. As expected, the harmonization potential varies across studies and variables. For example, the ‘‘age (years)’’ and ‘‘sex/gender (here, defined as female/male/other)’’ of the participant can be created by all studies, while “currently smoking tobacco (yes/no)” and “height (centimeters)” can be generated by eight of the 10 studies. Based on this use case, five studies (CoLaus|PsyCoLaus, Lc65+, SCQM, SKIPOGH, and SHeS-pp) out of the 10 can concurrently generate the following variables: ‘‘age (years)’’, ‘‘sex (female/male/other)’’, ‘‘highest level of education (compulsory education/upper secondary education/tertiary education)’’, ‘‘employment status (not currently employed/currently employed)’’ and ‘‘current tobacco use (yes/no)’’, offering the potential to co-analyse data from approximately 37 000 participants.

## What are the main strengths and weaknesses?

### Strengths

The SPHN-CC catalogue helps improve findability and serves as a key resource for optimizing the use of collected data. It facilitates exploring variable content and allows downloading Excel tables with selected lists of variables, which can be added to requests for access to data addressed to the studies. The SPHN-CC increases data interoperability not only between Swiss cohorts but also between Swiss and non-Swiss cohorts, thereby facilitating the building and consolidation of international networks and research consortia. Swiss cohorts have a long history of successful international collaborations[7–9]. However, each collaboration previously had to set up all necessary infrastructure independently, such as variable harmonization and documentation, without any centralised support. The SPHN-CC catalogue could greatly facilitate this type of exercise by providing a shared resource to streamline and support these collaborative efforts.

In addition to the SPHN-CC network, over 430 studies from 27 international networks are documented on the Maelstrom Research website. This allows users to easily search for information contributed not only by SPHN-CC but also by other international networks. The catalogue also facilitates the planning and organisation of research projects aimed at exploring temporal trends in selected fields, given that data collection spans decades in some cohorts. The SPHN-CC can also be used to highlight specific domains in which data collection gaps may exist, such as a detailed assessment of the multidimensionality of sex and gender or an in-depth exploration of professional exposures. This can be particularly useful for informing the implementation of new cohort studies and refining measures in ongoing ones. The catalogue can be used as a tool to explore similarities across cohorts and potential for impactful scientific collaborations, thereby guiding strategic decisions for future data collection.

### Weakness

The current list of cohorts featured in the network is relatively small, but the potential of the initiative will increase as more cohorts are added. One example is the Swiss Cohort Study on Lung and Heart Disease in Adults (SAPALDIA)[10], which, while not included, represents a significant data resource. SAPALDIA has established a publicly accessible and searchable meta-database, the Code Book Explorer, and has harmonized its data with international cohorts[10– 12]. Its phenotyping and questionnaire instruments, including tools for collecting medical history and medication data, served as the basis for the SHeS-pp study[13]. Accordingly, SAPALDIA data are largely harmonized with the SHeS-pp data currently described in the network. In some cases, study-specific data access may be limited, not allowing data transfer outside of the host institution or Switzerland. It may thus be required to base statistical analysis on individual participant data meta-analysis, aggregated data or use a federated approach like DataSHIELD[14,15]. The SPHN-CC facilitates the transition towards greater interoperability in the data of Swiss cohorts in general but is not a centralised data repository. Access to data from multiple studies remain a time-consuming process although standardization can significantly ease this process and led to fruitful collaborations in recent years. For example, SHCS not only developed the HICDEP protocol (https://hicdep.org/), which enabled efficient international data exchange and collaborative efforts, but also demonstrated the value of such collaborations in several joint research initiatives[16,17].

## Can I get hold of the data? Where can I find out more?

Data from the SPHN cohort consortium participating studies are not available through the catalogue. Access to individual participant data must be requested from the studies directly. Complete descriptions and information about the network and its individual studies and variables, as well as contact information for requesting access to data from each study, can be found here: https://mica.maelstrom-research.org/network/sphn-cc

**Supplementary Material 1:** Harmonization potential across partner studies for a selected ensemble of defined variables. Harmonization potential estimated based on the study-specific variables collected at baseline for 26 selected variables. The harmonization status is considered ‘‘complete’’ when study-specific variables could be used to generate the defined core variable, ‘‘partial’’ when it is possible but involve loss of information, and ‘‘impossible’’ when the core variable can’t be generated.

| Variables | Content | CoLaus-PsyCoLaus | Lc65+ | SCQM | SHCS | SIBDCS | SKIPOGH | SMSC | SSCS | STCS | SHES-pp |
| --- | --- | --- | --- | --- | --- | --- | --- | --- | --- | --- | --- |
| Age | Units: years | V | V | V | V | V | V | V | V | V | V |
| Sex | Categories: 1 = Female\|2 = Male\|3 = Other | V | V | V | V | V | V | V | V | V | V |
| Marital status | Categories: 1 = Single\|2 = Married/In a relationship\|3 = Divorced/Separated\|4 = Widowed | V | V | X | ~ | X | ~ | X | X | V | X |
| Highest level of education | Categories: 1 = Mandatory school\|2 = Other | V | V | V | V | X | V | X | V | V | V |
| Household monthly household income after taxes | Categories: 1 = < 4500 CHF\|2 = 4501-6000 CHF\|3 = 6001-9000 CHF\|4 = > 9000 CHF | X | ~ | X | X | X | X | X | X | V | V |
| Number of people in the household | Units: individuals | X | V | X | X | X | V | X | X | X | V |
| Ethnicity | Categories: 1 = Caucasian/White/European\|2 = African/Black\|3 = Asian/Pacific Islander\|4 = Other | ~ | X | X | V | X | V | V | V | V | X |
| Employment status | Categories: 0 = Not currently employed\|1 = Currently employed | V | V | V | X | X | V | X | X | ~ | V |
| Employment pattern | Categories: 1 = Part-Time\|2 = Full-Time | V | V | V | X | X | ~ | X | X | V | V |
| Ever smoked tobacco | Categories: 0 = No\|1 = Yes | V | V | V | V | X | V | X | X | V | V |
| Currently smoking tobacco | Categories: 0 = No\|1 = Yes | V | V | V | V | V | V | X | X | V | V |
| Currently drinking alcohol | Categories: 0 = No\|1 = Yes | V | V | V | V | X | V | X | X | X | V |
| Height - Measured or self-reported | Units: centimeters | V | V | V | V | V | V | V | X | X | V |
| Weight - Measured or self-reported | Units: kilograms | V | V | V | V | V | V | V | X | V | V |
| Heart rate at rest | Units: beats/min | V | V | X | X | X | V | X | X | X | V |
| Diastolic blood pressure | Units: mmHg | V | V | X | V | X | V | X | X | V | V |
| Systolic blood pressure | Units: mmHg | V | V | X | V | X | V | X | X | X | V |
| Occurrence of myocardial infarction | Categories: 0 = No\|1 = Yes | V | V | V | V | X | X | X | X | X | V |
| Occurrence of asthma | Categories: 0 = No\|1 = Yes | V | V | X | X | X | V | X | X | X | V |
| Occurrence of cancer | Categories: 0 = No\|1 = Yes | V | V | V | V | X | V | X | V | V | V |
| Occurrence of cerebrovascular disease | Categories: 0 = No\|1 = Yes | V | V | V | V | X | V | X | X | X | X |
| Occurrence of HIV infection | Categories: 0 = No\|1 = Yes | V | V | V | V | X | X | X | V | X | V |
| Occurrence of diabetes | Categories: 0 = No\|1 = Yes | V | V | V | V | V | V | X | X | X | V |
| Occurrence of high blood pressure | Categories: 0 = No\|1 = Yes | V | V | V | V | X | V | X | X | X | V |
| Occurrence of menopause | Categories: 0 = No\|1 = Yes | V | X | X | V | X | V | X | X | X | V |
| Total number of children | Units: individuals | V | V | V | X | X | X | X | X | X | V |
| V = Complete |  |  |  |  |  |  |  |  |  |  |  |
| X = Impossible |  |  |  |  |  |  |  |  |  |  |  |
| ~ = Partial |  |  |  |  |  |  |  |  |  |  |  |

## Data Availability

All data used in the present work is available online at: https://www.maelstrom-research.org/network/sphn-cc

## Ethics Approval

All participating studies received ethical approvals from their respective institutions and informed consent from participants. Investigators requiring access to study-specific data need to obtain ethical approval from their home institution and follow study-specific data access rules and procedures.

## Author contributions

MB, GH, MK, VP, NPH, JA, MB, IF, SOE initiated and designed the project. MB, IF, SOE were overseeing the project. The CoLaus-PsyCoLaus (MP, PV, PMV, JV, EC), Lc65+ (YH, JMB), SCQM (AS, CT), SHCS (HFG, KK), SHeS-pp (MB), SIBDCS (VP), SKIPOGH (MB, JPG), SMSC (JK, PB), SSCS (CC, CR) and STCS (MK) studies prepared and provided the metadata for the SPHN-CC. SEBT performed the harmonization potential analysis. JA, SEBT, MB, SOE, IF wrote the manuscript. All authors read and approved the manuscript.

## Conflicts of interest

The following co-authors declare no conflict of interest: AS, BP, CC, CR, CT, EC, IF, JA, JPG, JMB, JK, JV, KK, MB, MK, MI, MPRU, NPH, PB, PMV, PV, RW, SEBT, SOE, VP, YH H. F. G. has received research grants from the Swiss National Science Foundation, Swiss HIV Cohort Study, Yvonne Jacob Foundation, NIH, Gilead, ViiV, and is a subcontractor to a Bill and Melinda Gates foundation grant, paid to his institution; personal honoraria for data safety monitoring board or advisory board consultations from Merck, ViiV healthcare, Gilead Sciences, Janssen, Johnson and Johnson, Novartis, and GSK.

AR reports support to his institution for advisory boards and/or travel grants from MSD, Gilead Sciences, ViiV and Moderna, and an investigator initiated trial (IIT) grant from Gilead Sciences. All remuneration went to his home institution and not to AR personally, and all remuneration was provided outside the submitted work. MPRE has received unrestricted research grants from GlaxoSmithKline and the Swiss National Science Foundation.

## Fundings

This project was funded by the Swiss State Secretariat of Research and Innovation through the Swiss Personalized Health Network initiative. Each cohort provided in kind contributions.

SKIPOGH was supported by a grant from the Swiss National Science Foundation (FN 33CM30-124087).

The CoLaus|PsyCoLaus study was supported by unrestricted research grants from GlaxoSmithKline, the Faculty of Biology and Medicine of Lausanne, the Swiss National Science Foundation (grants 3200B0–105993, 3200B0-118308, 33CSCO-122661, 33CS30-139468, 33CS30-148401, 33CS30_177535, 3247730_204523, 320030_220190 and 324730_189130) and the Swiss Personalized Health Network (grant 2018DRI01).

The Swiss Multiple Sclerosis Cohort (SMSC) received funding from the Swiss Multiple Sclerosis Society and unrestricted grant funding from Biogen, Bristol Myers Squibb, Merck, Novartis, and Roche.

The Swiss HIV Cohort Study is financed by the by the Swiss National Science Foundation (grant #33FI-0_229621), and the SHCS research foundation. The data are gathered by the Five Swiss University Hospitals, two Cantonal Hospitals, 15 affiliated hospitals and 36 private physicians (listed in http://www.shcs.ch/180-health-care-providers).

The Lausanne cohort 65+ study (Lc65+) has been financed exclusively by public funds or non-profit organisations. Currently, funding is provided by the Public Health Department of canton Vaud, the Centre for Primary Care and Public Health (Unisanté) and by the Esther Locher-Gurtner Foundation.

The Swiss Systemic Lupus Erythematosus Cohort (SSCS) was supported by the Gebert-Rüf Foundation and the Fleurette Wagemakers Foundation and is currently financed by the SSCS Association and unrestricted research grants from GlaxoSmithKline, AstraZeneca and Otsuka.

The Swiss Inflammatory Bowel Cohort study (SIBDCS) was supported by the Swiss National Science Foundation (SNF-grant “Swiss IBD cohort study” N°33CS30-134274 (04.2011-03.2013), N°33CS30-148422 (04.2014-03.2018) and N°33CS30-177523 (04.2018-12.2021)

A list of rheumatology offices and hospitals contributing to the SCQM registry can be found at https://www.scqm.ch/en/about-scqm/active-institutions/. The SCQM thanks the patients for their participation in the registry. A list of current partners that support SCQM is available at https://www.scqm.ch/en/partners/. Partners from previous years can be found in the respective annual reports at https://www.scqm.ch/en/about-scqm/annual-reports/.

## Data availability

See the above section “Can I get hold of the data? Where can I find out more?” for more details.

## Use of artificial intelligence

Grammar proofreading was supported by ChatGPT-4 (OpenAI).

## Notes

### Author Declarations

The data source used for this work are entirely available on the network page this work describes: https://www.maelstrom-research.org/network/sphn-cc For the individual cohorts: CoLaus|PsyCoLaus: https://www.maelstrom-research.org/study/colaus-psycolaus-1 Lausanne Cohort 65+ (Lc65+): https://www.maelstrom-research.org/study/lc65 Swiss Clinical Quality Management in Rheumatic Diseases (SCQM): https://www.maelstrom-research.org/study/scqm Swiss HIV Cohort Study (SHCS): https://www.maelstrom-research.org/study/shcs Swiss Health Study - Pilot Phase (SHeS-pp): https://www.maelstrom-research.org/study/shes-pp Swiss Inflammatory Bowel Disease Cohort Study (SIBDCS): https://www.maelstrom-research.org/study/sibdcs Swiss Kidney Project on Genes in Hypertension (SKIPOGH): https://www.maelstrom-research.org/study/skipogh The Swiss Multiple Sclerosis Cohort-Study (SMSC): https://www.maelstrom-research.org/study/smsc Swiss Systemic Lupus Erythematosus Cohort Study (SSCS): https://www.maelstrom-research.org/study/sscs Swiss Transplant Cohort Study (STCS): https://www.maelstrom-research.org/study/stcs This work is a meta-analysis of already publicly available information from the catalogue. No additional data were used in the analysis—only the cohorts' metadata—and absolutely no sensitive or individual-level information is included.

### Summary of Updates

Added missing funding information for the Swiss Clinical Quality Management in Rheumatic Diseases (SCQM) cohort. Corrected few typographical errors and harmonised spelling throughout the manuscript according to UK English conventions.

